# Orang Asli Health and Lifeways Project (OA HeLP): Study Protocol

**DOI:** 10.1101/2021.10.24.21265442

**Authors:** Ian J. Wallace, Amanda J. Lea, Yvonne A. L. Lim, Steven K. W. Chow, Izandis bin Mohd Sayed, Romano Ngui, Mohd Tajudin Haji Shaffee, Kee-Seong Ng, Colin Nicholas, Vivek V. Venkataraman, Thomas S. Kraft

**Author notes:** These authors contributed equally (Ian J. Wallace, Amanda J. Lea). These authors contributed equally (Vivek V. Venkataraman, Thomas S. Kraft).

## Abstract

**Introduction:** Non-communicable disease (NCD) risk is influenced by environmental factors that are highly variable worldwide, yet prior research has focused mainly on high-income countries where most people are exposed to relatively homogenous and static environments. Understanding the scope and complexity of environmental influences on NCD risk around the globe requires more data from people living in diverse and changing environments. Our project will investigate the prevalence and environmental causes of NCDs among the indigenous peoples of Peninsular Malaysia, known collectively as the Orang Asli, who are currently undergoing varying degrees of lifestyle and sociocultural changes that are predicted to increase vulnerability to NCDs, particularly metabolic disorders and musculoskeletal degenerative diseases.

**Methods:** Biospecimen sampling and screening for a suite of NCDs (e.g., cardiovascular disease, type II diabetes, osteoarthritis, osteoporosis), combined with detailed ethnographic work to assess key lifestyle and sociocultural variables (e.g., diet, physical activity, technology usage), will take place in Orang Asli communities spanning a gradient from remote, traditional villages to acculturated, market-integrated urban areas. Analyses will, first, test for relationships between environmental variables, NCD risk factors, and NCD occurrence to investigate how environmental changes are affecting NCD susceptibility among the Orang Asli. Second, we will examine potential molecular and physiological mechanisms (e.g., epigenetics, systemic inflammation) that mediate environmental effects on health. Third, we will identify intrinsic (e.g., age, sex) and extrinsic (e.g., early life experiences) factors that predispose certain people to NCDs in the face of environmental change to better understand which Orang Asli individuals are at greatest risk of NCDs.

**Ethics:** Approval was obtained from multiple ethical review boards including a committee at the Malaysian Ministry of Health. This study follows established principles for ethical biomedical research among vulnerable indigenous communities, including fostering collaboration, building cultural competency, enhancing transparency, supporting capacity building, and disseminating research findings.

**Strengths and Limitations of This Study:**

- Environmental influences on non-communicable disease (NCD) risk are understudied outside of high-income countries, particularly among societies transitioning from traditional, non-industrial lifestyles to market-integrated, urban lifestyles.
- This multidisciplinary project aims to better understand how rapid lifestyle and sociocultural changes are affecting NCD risk among the indigenous peoples of Peninsular Malaysia, known collectively as the Orang Asli.
- More broadly, this project aims to provide insights useful for understanding the rising prevalence of NCDs in other low- and middle-income countries and societies experiencing rapid environmental changes.
- This project might be limited by the SARS-CoV-2 pandemic.

## INTRODUCTION

People today are getting sick and dying from a variety of non-communicable diseases (NCDs) that were much less prevalent among earlier generations. As recently as 100 years ago, the primary causes of morbidity and mortality in high-income countries (HICs) such as the United States were infectious diseases and malnutrition.^1^ In contrast, the greatest threats today are posed by NCDs that include metabolic disorders such as obesity, cardiovascular disease, and type II diabetes, musculoskeletal conditions such as osteoarthritis, back pain, and osteoporosis-related bone fractures, and neurological illnesses such as Alzheimer’s disease and other dementias.^2^ In recent decades, there have also been dramatic increases in the incidence and burden of NCDs in low- and middle-income countries (LMICs).^3^ Rates of obesity and related diseases, for example, are currently rising fastest in Asia and Latin America,^4-6^ and evidence suggests that even rural and subsistence-based societies in these regions are increasingly susceptible to NCDs.^6-10^ Such increases in NCDs are not necessarily associated with the same reductions in infectious diseases observed in HICs, leading to a “double burden of disease.”^11,12^ The escalating NCD pandemic is thus among the largest and most urgent global health concerns today.

NCD susceptibility is known to be influenced by intrinsic factors such as age, sex, and genetic variation, yet the rapid increase of NCDs within a few generations indicates that recent environmental changes strongly contribute to disease pathogenesis. Many NCDs thus appear to fit the definition of “mismatch diseases,” disorders caused by human bodies being inadequately or imperfectly adapted to novel features of modern environments.^13,14^ Nevertheless, explicitly testing the mismatch hypothesis, and more generally, identifying environmental factors responsible for the global NCD pandemic, has been challenging for three main reasons.

First, because most research to date has focused on HICs, the range of environments experienced by study participants is relatively static and homogenous.^15^ For example, essentially all people in HICs consume some amount of highly processed foods and rely regularly on labor-saving technologies. Thus, we are unable to test how the full spectrum of environmental variation—ranging from minimal exposure to full exposure—predicts NCD risk within a single population, which could be important for revealing nonlinear relationships as well as for increasing statistical power. Furthermore, people in HICs tend to experience similar environments throughout their lifetimes, making it difficult to disentangle the effects of exposures that occur in early life versus adulthood. Given the growing appreciation that early life conditions shape later life health and NCD risk,^16,17^ identifying study systems with more dynamic, within-lifetime variation is a major priority.

Second, environmental conditions—including lifestyle and sociocultural factors such as physical activity patterns, diet and food processing, technology usage, degree of market integration, social organization and stratification, gender relations, and divisions of labor—are highly variable among populations worldwide, so it is unlikely that findings from the Global North can be extrapolated to the rest of the world.^3,6^ Moreover, evidence suggests that both the types and impacts of environmental factors underlying NCD susceptibility may differ between populations. In the United States, for example, the environmental shifts believed to be most responsible for current high levels of obesity and associated NCDs are major declines in physical activity and greater consumption of energy-dense processed foods and drinks.^18-20^ Yet, outside of high-income countries, prevalence of metabolic NCDs is increasing even among societies in which most people are rarely able to avoid regular physical activity and/or have relatively little to moderate access to processed market foods.^8-10^ Also, people in many such societies have been documented to have a heightened sensitivity to obesity-related diseases for a given body mass index (BMI) compared to people in HICs,^21,22^ likely due to the extremely rapid rate of environmental changes occurring in LMICs.^23-25^ Ultimately, understanding the diversity and complexity of environmental influences on NCD risk worldwide, as well as preventing further increases in NCDs on a global scale, requires more data from people living in diverse environments.^15^

Third, previous attempts to understand the ultimate causes of NCDs through the lens of the mismatch hypothesis have relied on study designs that are not well suited for rigorously testing the idea. In particular, many studies have sought to test the mismatch hypothesis by comparing NCD prevalence and risk factors between HICs and small-scale, subsistence-level societies (e.g., hunter-gatherers, horticulturalists, pastoralists), which arguably have lifestyles that are more “matched” to their recent evolutionary history. These studies have repeatedly found low levels of NCDs such as obesity, cardiovascular disease, and type II diabetes in subsistence-level groups relative to HICs.^26-28^ This study design, however, confounds genetic background and recent evolutionary history with current lifestyle, limiting our ability to distinguish genetic versus environmental effects. A more robust study design would be to compare health between individuals engaging in traditional lifeways versus individuals from the same general genetic background living an industrial or post-industrial lifestyle. Such data are required for understanding how past selection regimes affect current NCD susceptibility.

Here, we present a research program that investigates the prevalence, underlying environmental causes, and evolutionary explanations for NCDs among the indigenous peoples of Peninsular Malaysia, known collectively as the Orang Asli. The Orang Asli (meaning “original people” in Malay) occupy a relatively small geographic area but are exposed to a wide range of environmental conditions. Traditionally, the Orang Asli live in relatively small, remote forest camps and villages and subsist on some combination of hunting, fishing, wild food collection, horticulture, and trade of forest products.^29,30^ Today, however, no Orang Asli communities are completely isolated from outside economic and cultural influences due to the rapid expansion of industries, governmental regulation, the market economy, and urban areas across Malaysia over the last half-century.^31-38^ As a result, Orang Asli groups have been experiencing varying types and degrees of environmental change (e.g., ecological degradation, acculturation, market integration, urbanization) that are occurring rapidly and within the span of an individual lifetime. At one extreme, some Orang Asli communities remain relatively isolated and continue to adhere largely to traditional lifestyles, but at the other extreme, some communities are now bound by urban areas, highly acculturated, and fully integrated into the market economy. Not surprisingly, while historical data suggest that NCDs were once rare among the Orang Asli,^39-44^ recent work indicates that NCDs are a growing problem, especially in the more urbanized, acculturated, and market-integrated communities.^45-49^ Infectious diseases also continue to be major health threats for the Orang Asli.^50-53^

Our project—called the Orang Asli Health and Lifeways Project (OA HeLP)—is a collaboration between anthropologists, biomedical researchers, clinicians, and study communities that aims to better understand how and why environmental changes are occurring among the Orang Asli and how such changes are affecting individuals’ susceptibility to NCDs. Presently, our focus is on metabolic disorders (obesity, cardiovascular disease, type II diabetes) and musculoskeletal degenerative diseases (osteoarthritis, osteoporosis), though research on additional types of NCDs is expected in the future. Specifically, the primary goals of the project are to: (1) conduct ethnographic fieldwork in Orang Asli communities to collect data on rapidly changing environmental variables including lifestyle and sociocultural conditions; (2) conduct biomedical screening to collect anthropometric, imaging, and physiological data pertaining to general health, NCD risk, and NCD diagnosis; (3) test for relationships between environmental variables, NCD risk factors, and NCD occurrence to investigate how environmental changes are affecting NCD vulnerability among the Orang Asli; (4) understand the molecular and physiological mechanisms (e.g., epigenetics, systemic inflammation, infectious diseases) that mediate environmental effects on health; and (5) identify intrinsic (e.g., age, sex) and extrinsic (e.g., early life experiences) factors that predispose certain people to NCDs in the face of environmental change, in order to understand which Orang Asli individuals are at greatest risk of NCDs. More broadly, this project aims to provide insights useful for understanding the rising prevalence and burden of NCDs in other populations and regions of the world.

## METHODS AND ANALYSES

### Study Population and Setting

The population of Peninsular Malaysia is multiethnic, with the majority people being ethnically Malay and predominantly Muslim, and with large minorities of Malaysian people of Chinese and Indian ancestry. The Orang Asli comprise less than 1% of the population (approximately 200,000 people) and include at least 19 distinct ethnolinguistic groups, which are typically divided into three broad categories (figure 1): the Semang (traditionally, speakers of northern Aslian languages and primarily nomadic hunter-gatherers), Senoi (traditionally, speakers of central Aslian languages and primarily horticulturalists), and Aboriginal Malay (traditionally, speakers of Melanesian language dialects and practitioners of mixed subsistence and cash crop economies).^29,30^ The genetic history of the Orang Asli is complex and an area of active study. The three broad groups (Semang, Senoi, and Aboriginal Malay) are genetically distinguishable, though they are generally more similar to one another than they are to other surrounding Asian populations.^54^ Currently, our project is focused on six particular ethnolinguistic groups—the Batek, Jahai, Semai, Temiar, Temuan, and Jakun—which represent two examples each of Semang, Senoi, and Aboriginal Malay peoples, respectively. Additional groups are expected to be added to the study in the future. Study communities are located in different areas of Peninsular Malaysia and span a gradient from remote, traditional camps and villages to acculturated, market-integrated urban areas (figure 2).

**Figure 1.**
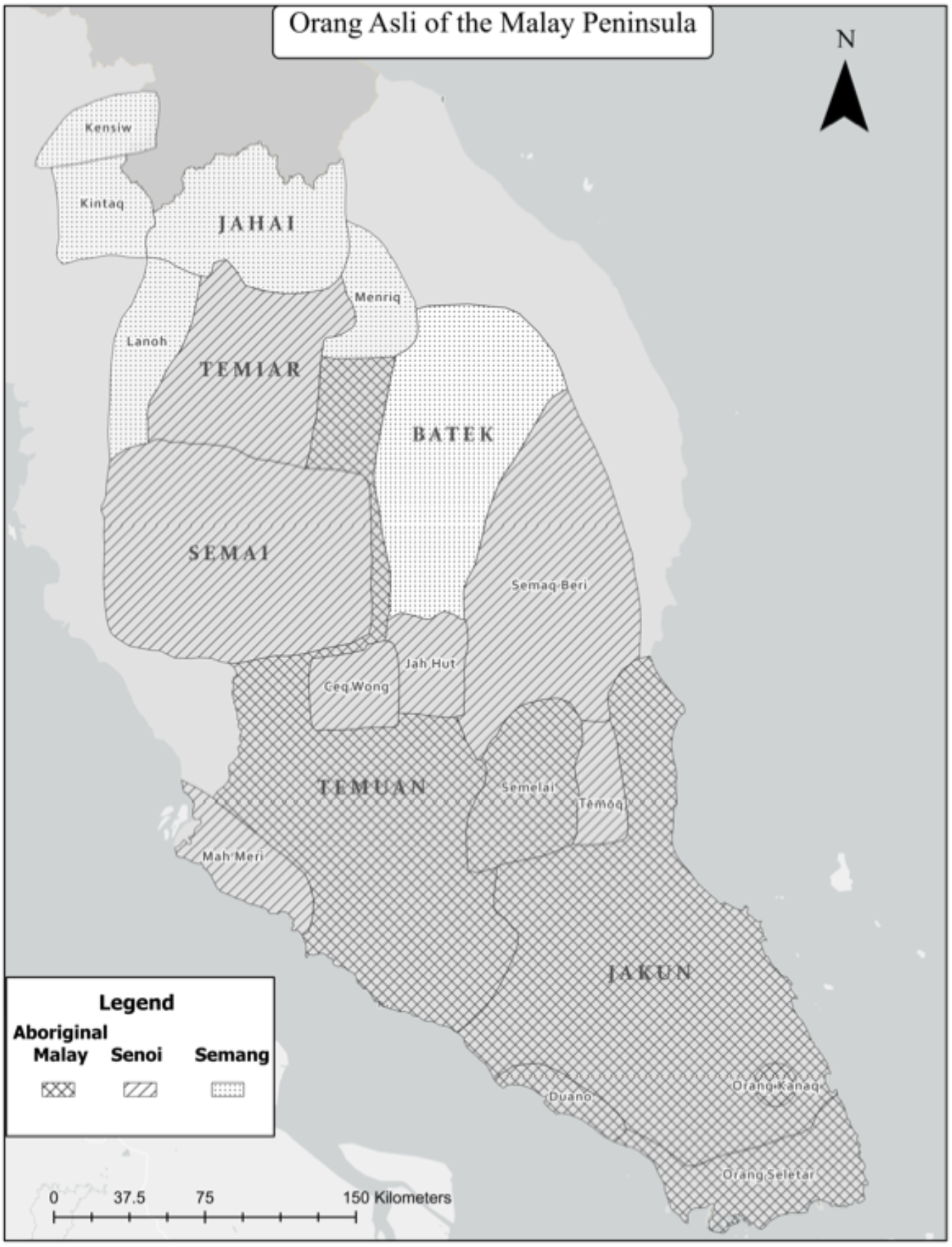
Map of Peninsular Malaysia showing the approximate locations of Orang Asli groups.

**Figure 2.**
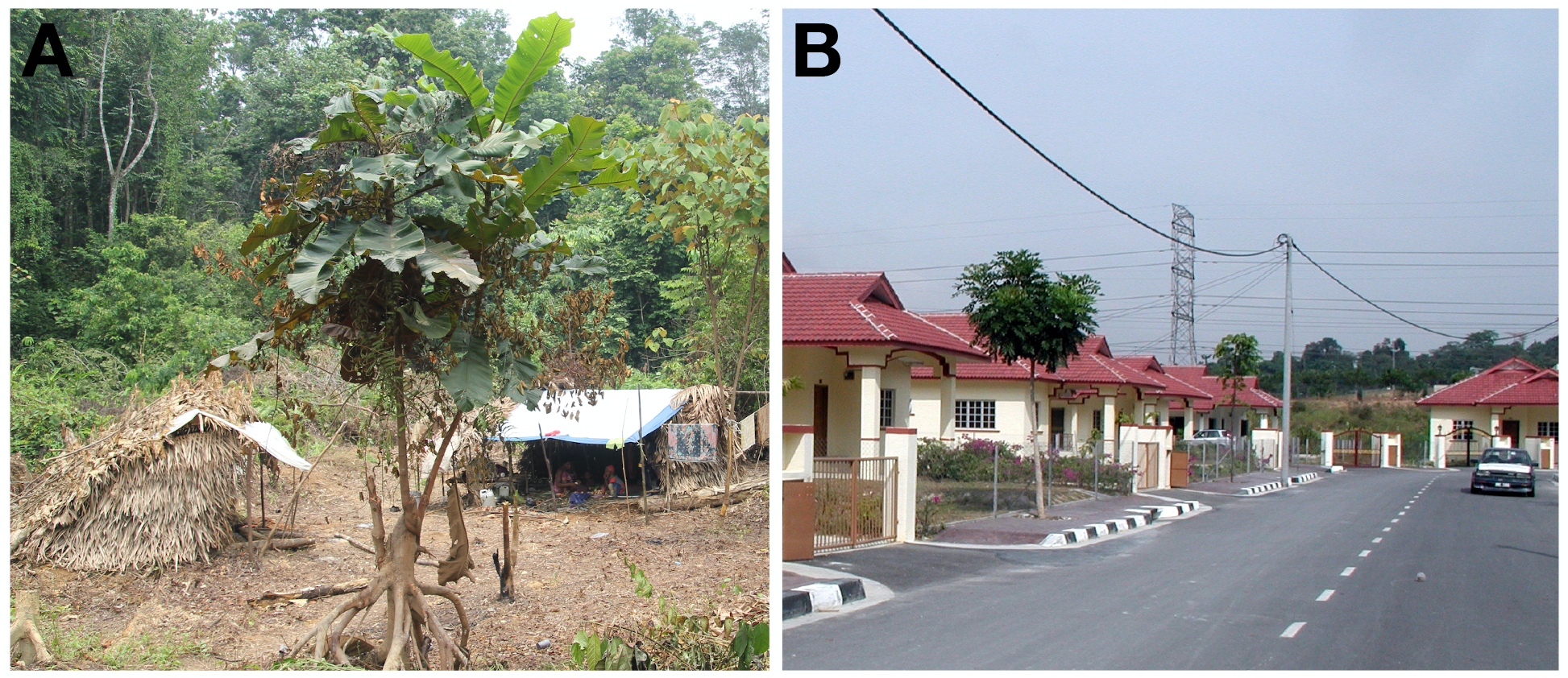
Orang Asli study communities span a gradient from (A) remote, traditional camps and villages to (B) acculturated, market-integrated urban areas.

Among the environmental changes experienced by the Orang Asli over the last half-century, two trends deserve special mention because their effects on lifestyles and sociocultural conditions have been pronounced. First, a key component of efforts to accelerate growth of the national market economy has been the expansion of industries focused on plantation agriculture (particularly oil palm and rubber) and natural resource extraction (particularly timber, tin, and petroleum).^55-57^ As a result, Malaysia has experienced rapid, marked deforestation,^58-60^ which has fragmented and altered large fractions of the lands traditionally occupied by the Orang Asli and has had broad negative impacts on biodiversity and water, soil, and air quality.^61-65^ Dispossessed of the land and resource base necessary for traditional subsistence strategies (most Orang Asli do not hold official title to their traditional lands), many communities have shifted their livelihoods to include wage labor, often in the very industries responsible for their land and resource disposession.^33^ Second, a longstanding objective of the Malaysian government has been to promote the assimilation of the Orang Asli into mainstream Malaysian society and the integration of Orang Asli economies with the national market economy.^30-32^ To this end, government programs have been established to resettle and regroup the Orang Asli into consolidated communities with modern facilities including administrative centers, schools, shops, clinics, houses, and roadways.^33^ In addition, community members are typically provided some form of income-generating activity (e.g., rubber or oil palm). Ultimately, due to these two trends, many Orang Asli currently live in highly acculturated, market-integrated contexts.^33,35-37^ Variation exists, however, in how long people have lived in such contexts, with some people having transitioned only recently and others having lived in such contexts for their entire lives.

Notwithstanding the strength of industry, government, and other forces, many Orang Asli continue to adhere largely to traditional lifestyles, albeit on lands that are threatened or degraded relative to those occupied by earlier generations. The most averse to change have been Semang groups (e.g., Batek, Jahai), which traditionally practice a hunter-gatherer lifestyle.^31,34,66^ Because of the traditionally nomadic, egalitarian, and autonomous nature of these groups,^66-68^ most have resisted sedentarization and the authority of industry, government, and other entities.^31,66^ Consequently, there still exist remote communities that rely heavily on the availability of natural resources.

### Study Design

Our current project uses a cross-sectional cohort design^69^ that will be implemented over at least five years. Participant recruitment has begun, and pilot data have been collected. However, due to the SARS-CoV-2 pandemic, it has not yet been possible to implement our complete protocol. Our expectation is that the project will be fully operational by 2022 or 2023.

Research in Orang Asli communities will occur in two phases meant to assess lifestyle/sociocultural conditions and NCD susceptibility, respectively. In the first phase, ethnographic fieldwork will be carried out by anthropologists aided by community members to conduct interviews, administer questionnaires, construct demographic profiles, and collect data on key lifestyle and sociocultural variables, as well as physical activity. Depending on the size of the community, this phase will take 2-6 weeks to complete. In the second phase, biomedical researchers will join anthropologists to carry out biomedical screening to collect anthropometric, imaging, and physiological data pertaining to general health, NCD risk, and NCD diagnosis, particularly metabolic and musculoskeletal NCDs. In each community, this phase will take 3-7 days to complete.

Biomedical screening will be conducted in partnership with mobile clinics organized by the Federation of Private Medical Practitioners’ Associations of Malaysia (FPMPAM). The FPMPAM is the national body representing doctors in private practice in Malaysia. Among other programs, the FPMPAM organizes mobile clinics that travel to Orang Asli communities throughout Peninsular Malaysia to perform routine physical examinations, provide primary care, and arrange for transport to a hospital when necessary (a program known as Drs4All, led by S.K.W.C.). These mobile clinics play a critical role in providing health care to the Orang Asli because (1) many communities are located far from clinics and hospitals, and (2) even in communities that have clinics, the clinics tend to be understaffed and lack supplies and medicines.^50^

There are multiple benefits to scheduling biomedical screening to coincide with mobile clinic visits. First, the overall safety and comfort of the participants will be enhanced by having physicians nearby. Second, based on many positive prior interactions between Orang Asli communities and the mobile clinics, community members trust the medical expertise of members of the mobile clinics, whom they can consult if they have any questions or concerns about participating in biomedical screening. Third, biomedical researchers will be collecting specimens and data that could be useful to physicians in diagnosing medical conditions. Thus, the project aims to improve equitability between researchers and study participants by directly providing tangible benefits to study communities.

### Study Sample and Recruitment

All people >18 years old who self-identify as ethnically Orang Asli are eligible to participate in the study. However, people <40 years old, as well as pregnant woman, will be excluded from radiographic musculoskeletal imaging (see below). Participation in the study will not affect whether people are able to receive care from the mobile clinics.

The following formula was used to estimate the target sample sizes for the study:

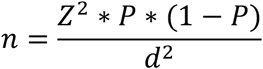

where n is sample size, Z is the statistic corresponding to level of confidence, P is expected prevalence, and d is precision/margin of error.^70^ We assumed standard parameters for Z (specifically, a 99% confidence level) and a prevalence of 0.5 (the recommended value for diseases whose prevalence is not known *a priori*, and therefore the value that captures the largest range of possibilities). Based on these parameter values, we will aim for a sample size of 1000 people, approximately half women and half men, to achieve a <5% margin of error when estimating the prevalence of any given NCD.

Appropriate and willing Orang Asli communities to be included in the project have been identified through existing relationships developed over many years of prior work by multiple members of our team. Our process of recruitment at the community level involves two general steps, which are then followed by the individual-level consent process. First, permission to conduct research is sought from community leaders. Traditionally, the existence of “leaders” varies across Orang Asli groups, but today most communities have individuals who maintain official titles of *penghulu* or *tok batin* assigned by the government that indicate a leadership role. In more egalitarian communities, initial meetings are held with an advisory committee of community members. Second, researchers and community leaders hold public meetings to discuss the project, including research questions and goals, methods and procedures, participant inclusion and exclusion criteria, and participant compensation (usually in the form of food, clothing, tools, and other staples).

### Study Procedures: Ethnographic Fieldwork

#### Interviews and questionnaires

Interviews will be conducted, and questionnaires administered, to construct demographic profiles and collect data on key lifestyle and sociocultural domains, including location and environmental surroundings, subsistence practices, diet and food processing, access to education and modern technology, degree of market integration, social organization and stratification, gender relations, divisions of labor, and early life environmental conditions. Questionnaire data will be summarized in two tiers. In the first tier, we will combine questions within each of the domains of interest to create composite indices of participant lifestyles that span a gradient from “traditional” to “non-traditional.” Continuous quantitative variables will be polarized (so that high values represent “non-traditional” lifestyles), standardized (e.g., to a standard normal scale), and summed to produce a score for each domain. In the second tier, we will incorporate information from all domains to create weighted composite scores (such that each domain carries the same weight).

In addition to characterizing lifestyle during interviews, we will also ask participants to complete a questionnaire about their perceived health, which will be comprised of questions translated into Malay from the Patient-Reported Outcomes Measurement Information System (PROMIS).^71^ Included are questions about musculoskeletal pain and disability, as well as other aspects of physical and mental health.

#### Measurements of physical activity

Accelerometry will be used to objectively measure physical activity, which is a documented risk factor for NDCs including obesity, cardiovascular disease, type II diabetes, osteoarthritis, and osteoporosis.^72-76^ Participants will be given Axivity accelerometers and asked to wear them for 5-7 days.^77^ These waterproof units weigh little (11g), thus causing no inconvenience to the participant. Accelerometry data will then be processed to quantify variables such as (1) overall physical activity levels (e.g., step counts, time spent in moderate-to-vigorous activities), (2) gait characteristics (e.g., step length, voluntary walking speed) and stability, and (3) postural behaviors (e.g., knee-bending activities).^77-79^

### Study Procedures: Biomedical Screening

#### Anthropometry

Anthropometric data will be collected to measure BMI and adiposity and diagnose obesity, which are documented risk factors for NDCs including cardiovascular disease, type II diabetes, and osteoarthritis.^80-83^ Standing height will be measured with a stadiometer. Body weight and body fat percentage will be measured with a digital bioelectrical impedance scale. Waist circumference will be measured with a tape measure.

#### Blood pressure

A stethoscope and arm cuff will be used to measure resting blood pressure and diagnose hypertension, which is a documented risk factor for many NCDs including cardiovascular disease and osteoarthritis.^75,84^ Participants will be resting in a seated position for at least 10 min before measurement, and three measurements will be taken 1 min apart.

#### Blood samples

Blood samples will be collected to assess biomarkers of lipid and glucose metabolism and chronic low-grade systemic inflammation, which are documented risk factors for NDCs including cardiovascular disease, type II diabetes, and osteoarthritis.^75,85-88^ Blood samples will be preserved on: (1) Whatman 903 Protein Saver Cards to study circulating biomarkers of inflammation (e.g., C-reactive protein, IL-6) and (2) Whatman FTA cards to study genome-wide DNA methylation levels in the context of lifestyle effects on gene regulation. Blood will also be used with multiple point-of-care devices including: (1) a CardioCheck Plus Analyzer to measure levels of blood lipids and glucose and to diagnose dyslipidemia; (2) a Hemocue WBC DIFF system to measure white blood cell counts; and (3) an A1CNow+ system to measure percent of glycated hemoglobin levels and diagnose type II diabetes.

#### Fecal and urine samples

Fecal samples will be collected to analyze the gut microbiome and intestinal parasites, which are known mediators of chronic low-grade systemic inflammation and hence potentially NCDs including cardiovascular disease, type II diabetes, and osteoarthritis.^89-91^ Urine samples will be collected to analyze biomarkers of bone and cartilage turnover (e.g., crosslinked C-(CTX) and N-(NTX) telopeptides of type I collagen).^92,93^ Participants will be given two opaque disposable plastic cups with lids in the evening and asked to collect the next day’s first feces and urination and return them to the biomedical team.

#### Bone ultrasound

Quantitative ultrasonography images will be collected from participants to measure bone mineral density and diagnose osteoporosis.^94^ Quantitative ultrasonography of the radius and tibia will be conducted with a portable MiniOmni probe. Probes will be placed at the medial aspect of the distal one-third of the radius of the non-dominant arm and at the anteromedial aspect of the midshaft tibia of the left leg.

#### Hand photos

A digital photo of the dorsum of both hands will be collected to diagnose osteoarthritis in the hand joints.^95^ The participant will relax their hands and place them palm down on a plastic grid pad, with fingers apart and thumbs angled approximately 30°.

#### Knee x-rays

Among people aged 40 years and older, we will collect knee radiographs to diagnose knee osteoarthritis. These will be collected using a portable MinXray Impact digital x-ray system, along with a SynaFlexer frame to standardize the positioning of participants’ knees in fixed-flexion view.^96^ The overall effective dose for each knee x-ray will be approximately 0.001 milliSieverts, which is equivalent to roughly 3 hours of natural background radiation.^97^

#### Knee pain sensitivity tests

Among people aged 40 years and older, we will also measure knee pain sensitivity related to osteoarthritis using standardized quantitative sensory testing.^98^ Specifically, pressure pain threshold will be measured using algometry to assess sensitivity to pain evoked by mechanical stimulation of nociceptors. Pain pressure threshold will be assessed by applying an algometer (1 cm^2^ rubber tip) to each patella at a rate of 0.5 kg/sec and recording the level at which the participant states that the pressure first changed to slight pain.

### Data Analysis

#### Overview

Our primary goals for data analysis are to: (1) estimate the prevalence of metabolic and musculoskeletal NCDs among the Orang Asli; (2) test for relationships between environmental variables, NCD risk factors, and NCD occurrence; (3) examine molecular and physiological mechanisms that mediate environmental effects on health; and (4) identify intrinsic and extrinsic factors that predispose certain people to NCDs in the face of environmental exposures. Below, we provide an overview of the specific strategies we will use to achieve these goals.

#### Estimating NCD prevalence

Anthropometric, imaging, and physiological data will be used to determine the presence or absence of the following NCDs in each participant: (1) overweight (25≤BMI<30) and obesity (BMI≥30); (2) hypertension (blood pressure>135/85); (3) type 2 diabetes (HbA1c of 6.5% or higher) and prediabetes (HbA1c of 5.7-6.4%); (4) metabolic syndrome (diagnosed using of combination of data on waist circumference, blood pressure, and blood lipid and glucose levels); (5) knee and hand OA (diagnosed from radiographs and photos by qualified experts); and (6) osteoporosis (T-score of −2.5 or below) and osteopenia (T-score between −1.0 and −2.5). These data will be used to estimate the prevalence of each NCD, which will be compared to estimates from other studies including those from HICs.

#### Testing for environmental effects on NCD risk and occurrence

To assess the effects of environmental variables on NCD risk and occurrence, we will first derive several composite measures of lifestyle from interviews and questionnaires, as described above. We will then use linear and generalized linear mixed models controlling for key covariates (e.g., age, sex) to ask whether specific environmental variables or our composite indices predict continuous variables associated with NCD risk, as well as binary data on NCD occurrence (presence/absence). Continuous variables of interest include BMI, waist circumference, body fat percentage, blood pressure, blood lipid and glucose levels, systemic inflammation, gut microbiome composition, intestinal parasite load, knee pressure pain threshold, levels of bone and cartilage turnover biomarkers, bone mineral density, and physical activity. In addition to examining the effects of individual environmental variables and using composite measures to reduce the dimensionality of the interview data, we will also apply techniques that are well suited to infer complex relationships between multiple predictor variables and an outcome, such as random forests, LASSO, or structural equation modeling.

#### Examining the mechanistic basis of environmental effects on NCD risk and occurrence

Several candidate molecular and physiological mechanisms will be considered that are known to be environmentally responsive and involved in the etiology of NCDs^99-102^, including: (1) genome-wide DNA methylation, an epigenetic gene regulatory mechanism; (2) IL-6 and CRP, circulating biomarkers of systemic inflammation; and (3) proportional and total counts of neutrophils, lymphocytes, basophils, eosinophils, and monocytes, which provide information about immune function and current infection status. To assess the involvement of these and other candidate mechanisms in mediating environmental effects on NCD risk factors and occurrence, we will first ask whether any of our environmental variables (composite or individually, focusing on those that are identified as most predictive of NCD risk factors and occurrence) predict variation in our mechanistic variables (using methods appropriate for each data type, e.g., references 103 and 104). Then, we will use formal mediation analyses^105,106^ to assess causal relationships between environmental variables, mechanisms, and NCD risk factors and occurrence, and to estimate the proportion of the total effect explained by a given mediator.

#### Examining inter-individual variation in NCD risk and occurrence

In all likelihood, environmental effects on NCD risk and occurrence will vary among individuals. A goal of our project is to identify factors that predispose individuals toward susceptibility versus resilience in the face of environmental change. To this end, we will test the degree to which several intrinsic and extrinsic factors mediate environmental influences on NCD risk and occurrence, including age, sex, and early life experiences.^16,17^ To do so, we will use linear and generalized linear mixed models to test for interaction effects for all environmental and NCD-related outcome variable combinations that were found to be significant in previous analyses. Analyses will control for multiple hypothesis testing using a Storey-Tibshirani false discovery rate approach^107^, and all code will be made publicly available on Github.

### ETHICS

Procedures for this study have been reviewed and approved by the Medical Review and Ethics Committee of the Malaysian Ministry of Health (protocol ID: NMRR-20-2214-55565) and the Institutional Review Board of the University of New Mexico in the United States (protocol ID: 14420). Informed consent will be obtained and documented for all study participants. All data obtained in the study will be kept confidential, securely protected and managed, and used only for research purposes. Throughout the project, we will follow established principles for ethical biomedical research among indigenous communities, including fostering collaboration, building cultural competency, being transparent about research practices, supporting capacity building, and disseminating research findings.^108^

### Participant and Public Involvement

In 2020, we hosted a multi-day public webinar series in collaboration with the Orang Asli Archive at Keene State College in the United States entitled “Orang Asli Health and Well-Being.” The series brought together a large international group of anthropologists, biomedical researchers, physicians, activists, and members of the general public, including many Orang Asli, to identify the greatest health challenges faced by the Orang Asli today. There emerged a strong consensus that NCDs are a rapidly escalating problem and that lifestyle changes are almost certainly largely to blame. These conclusions were consistent with observations made by many of us through our work in different domains. One of us (I.b.M.S.) is Orang Asli and the Director of the Hospital Orang Asli, Gombak, a facility devoted exclusively to the care of Orang Asli that has experienced a growing number of patients with NCDs in recent years. Physicians involved in the FPMPAM mobile clinics (S.K.W.C. and M.T.H.S.), as well as Malaysian biomedical researchers (Y.A.L.L., R.N., and K.-S.N.), have noted similar trends in the Orang Asli communities in which we work. Another author (C.N.) directs the Center for Orang Asli Concerns, a human rights organization that advocates for the Orang Asli, and through this work has received an increasing number of reports of concerns from Orang Asli about NCDs. Ultimately, it was insights gleaned from diverse perspectives in the webinar series, together with our own observations, that motivated the current aims, study design, and methodology of OA HeLP.

We are committed to carrying out this project as a collaboration with the Orang Asli communities in which we work. This collaboration will take at least four forms. First, local community leaders and members will be consulted regarding all aspects of the research process including the questions and goals of the project, study procedures, participant compensation, data analysis and management, and results dissemination. During fieldwork, these consultations will occur with community leaders on a continuous basis and with community members during regular public meetings. During non-fieldwork periods, consultations will continue through non-research-related follow-up visits to study communities, email and text messaging, and videoconferencing. As a result of these consultations, we fully expect the scope of OA HeLP to eventually expand to include additional research questions and goals. We also expect our current questions and goals to be enhanced by the perspectives of community members, and that our protocol will warrant some adjustment to be better suited for certain communities. Second, in study communities, we will employ as many Orang Asli as possible to participate in conducting interviews, collecting data, translation, community relations, and more. These individuals will be especially well positioned to provide input on the effectiveness and cultural appropriateness of our fieldwork as it occurs. Third, we aim to recruit and provide funding for Orang Asli graduate students and post-doctoral researchers to join the OA HeLP team and use OA HeLP resources and infrastructure to pursue their own health-related research goals in study communities. Our project will also support scientific training among Orang Asli from study communities through a program organized by the FPMPAM, in which Orang Asli individuals receive funding to work at private clinics across Malaysia where they receive education in first aid and emergency medicine before returning to their communities where they then serve as medical “first responders.” Fourth, all individuals in Orang Asli study communities who make substantial contributions to the research program will be invited to be co-authors on scientific publications arising from the research.

### Plan for Results Dissemination

A critical part of OA HeLP is the communication of our findings to study communities. As results become available, they will be presented in layperson’s terms at public meetings held in each study community, during which community members will be able to ask questions and provide their own perspectives on the findings. These meetings will provide important opportunities for developing interpretations of results that incorporate Orang Asli viewpoints. During meetings, we will discuss the prevalence of various NCDs and risk factors for those diseases, as well as the lifestyle factors that our research indicates are most closely associated with those NCDs. We will also discuss potential lifestyle adjustments that individuals could make to lower their vulnerability to NCDs. In addition, we will present our findings to the Malaysian Ministry of Health in written reports and in-person presentations. The Malaysian Ministry of Health oversees all government-run public health and prevention services for the Orang Asli. Thus, communicating our findings to the Malaysian Ministry of Health has the potential to improve health care among the Orang Asli by generating data that could be used to develop and provide more effective health services.

To publicize OA HeLP more broadly, we will maintain a website (www.orangaslihealth.org) that summarizes our progress and findings and provides photos from our fieldwork. Also, we will hold a public webinar series to present our finding in layperson’s terms, which will be advertised through our universities, the FPMPAM, Center for Orang Asli Concerns, and various social media outlets. In addition to communicating results to study communities and the general public, our findings will be presented to the scientific community in peer-reviewed journal articles and at national and international scientific conferences. Whenever possible, we will publish in open access formats. Upon request, anonymized data will be made available to other researchers pending data-use agreements and approval from a review committee made up of ourselves and Orang Asli community leaders. We welcome researchers to contact us if they are interested in becoming involved in OA HeLP, particularly researchers in Malaysia who share our research interests and are committed to improving Orang Asli health.

## Data Availability

All data produced in the present study are available upon reasonable request to the authors

## Acknowledgements

For critical insights into Orang Asli health and for helping motivate this project, we thank the participants of the 2020 public webinar entitled “Orang Asli Health and Well-Being.” For helpful advice in developing the study protocol, we thank Drs. Silvia Del Din, Orrin Devinsky, Karen Endicott, Kirk Endicott, David Felson, Michael Gurven, Brian Hainline, Nicholas Holowka, Helgi Jónsson, Tuhina Neogi, Maude Phipps, and Lynn Rochester. We are also deeply grateful to the Orang Asli communities that are collaborating on this project.

## Authors’ contributions

IJW, AJL, YALL, SKWC, IbMS, VVV, and TSK conceived of the study. All authors contributed to the development of the study protocol. IJW, AJL, VVV, and TSK drafted the manuscript, and YALL, SKWC, IbMS, RN, MTHS, K-SN, and CN edited the manuscript.

## Funding

OA HeLP is currently funded by the United States National Institutes of Health (subcontract of P30AR072571), University of New Mexico, Vanderbilt University, University of Calgary, and University of Utah.

## Competing interests

The authors declare that they have no competing interests.

## Patient consent for publication

Not required.

## Notes

### Competing Interest Statement

The authors have declared no competing interest.

### Author Declarations

Medical Review & Ethics Committee (MREC) of the Ministry of Health Malaysia gave ethical approval for this work. The Institutional Review Board of the University of New Mexico gave ethical approval for this work.

